# Deep Learning for Breast Mass Discrimination: Integration of B-Mode Ultrasound & Nakagami Imaging with Automatic Lesion Segmentation

**DOI:** 10.1101/2025.09.14.25335722

**Authors:** Walid Hassan, Murad Hossain

## Abstract

**Objective:** This study aims to enhance breast cancer diagnosis by developing an automated deep learning framework for real-time, quantitative ultrasound imaging. Breast cancer is the second leading cause of cancer-related deaths among women, and early detection is crucial for improving survival rates. Conventional ultrasound, valued for its non-invasive nature and real-time capability, is limited by qualitative assessments and inter-observer variability. Quantitative ultrasound (QUS) methods, including Nakagami imaging—which models the statistical distribution of backscattered signals and lesion morphology—present an opportunity for more objective analysis.

**Methods:** The proposed framework integrates three convolutional neural networks (CNNs): (1) NakaSynthNet, synthesizing quantitative Nakagami parameter images from B-mode ultrasound; (2) SegmentNet, enabling automated lesion segmentation; and (3) FeatureNet, which combines anatomical and statistical features for classifying lesions as benign or malignant. Training utilized a diverse dataset of 110,247 images, comprising clinical B-mode scans and various simulated examples (fruit, mammographic lesions, digital phantoms). Quantitative performance was evaluated using mean squared error (MSE), structural similarity index (SSIM), segmentation accuracy, sensitivity, specificity, and area under the curve (AUC).

**Results:** NakaSynthNet achieved real-time synthesis at 21 frames/s, with MSE of 0.09% and SSIM of 98%. SegmentNet reached 98.4% accuracy, and FeatureNet delivered 96.7% overall classification accuracy, 93% sensitivity, 98% specificity, and an AUC of 98%.

**Conclusion:** The proposed multi-parametric deep learning pipeline enables accurate, real-time breast cancer diagnosis from ultrasound data using objective quantitative imaging.

**Significance:** This framework advances the clinical utility of ultrasound by reducing subjectivity and providing robust, multi-parametric information for improved breast cancer detection.

## I. INTRODUCTION

Breast cancer is the most frequently diagnosed malignancy in women [1] and ranks second only to lung cancer in global cancer-related mortality [2]. To optimize breast cancer treatment and improve survival rates, early detection and precise diagnosis of breast cancer are vital. [3]. Recent studies have demonstrated that early detection of breast cancer through routine screening significantly improves survival rates. For instance, breast cancer patients in the US in stage I over 40 years old showed a 10-year relative survival rate higher than disease-specific survival, even exceeding 100%, indicating better survival than individuals of the same age without cancer [4]. While mammography serves as a primary screening tool for breast cancer [5], its limitations in dense breast tissue have prompted the integration of ultrasound imaging as a complementary modality [6]. The sensitivity and specificity of mammography for breast cancer detection were 62% to 86% and 66% to 91% with 95% CI, respectively, highlighting lower diagnostic performance compared to ultrasound 87% to 97% and 74% to 91% with 95% CI, respectively [6]. Ultrasound imaging is a non-invasive, portable, real-time, and cost-effective modality, making it a valuable addition to screening protocols [7].

However, the high variability in the diagnostic performance of ultrasound imaging is often limited by operator dependency [8], interpretive variability [8], and a lack of quantitative information [9]. These limitations motivate the researchers to develop advanced computational techniques for incorporating quantitative parameters from ultrasound imaging to enhance diagnostic capabilities [9]. Quantitative ultrasound imaging (QUS) enhances diagnostic capabilities by extracting parameters, such as the speed of sound, attenuation, and backscatter coefficients, from ultrasound radiofrequency (RF) data, providing objective tissue characterization [10]. Nakagami, a QUS parameter, quantifies ultrasound backscatter statistics, with studies demonstrating its ability to differentiate malignant and benign tumors. Previous research showed that Nakagami parametric imaging outperformed standard B-mode statistics, achieving 92% sensitivity, 72% specificity, and an AUC of 81%. Malignant tumors exhibited significantly lower mean Nakagami shape parameters (0.55 ± 0.12) than benign lesions (0.69 ± 0.12), highlighting the method’s superior capability to capture microstructural heterogeneity compared to B-mode’s performance, which had 60% sensitivity and 72% specificity. [11]. Recent advancements in integrating texture analysis with Nakagami parametric images have further improved performance, achieving 90.38% sensitivity and 89.58% specificity with an AUC of 94% when combining texture features and mean pixel values from tumor cores and peritumoral margins [12]. These developments highlight Nakagami imaging’s potential as a complementary tool to B-mode ultrasound, offering quantitative insights into tumor microstructure. For example, using a convolutional neural network (CNN) method, Muhtadi et al. achieved a detection accuracy of 87% with a combined Nakagami and B-mode envelope, compared to 82% when using B-mode alone [13]. However, a limitation of this study is the limited clinical data available for training and testing the CNN method, which consisted of only a 230-patient dataset comprising 152 benign and 78 malignant cases. While Nakagami imaging has shown promise in tissue characterization, it remains primarily used in research settings. It has not been adopted as a standard feature in diagnostic ultrasound systems due to the high computational cost. Additionally, typical B-mode images from a clinical ultrasound system cannot be used to generate Nakagami images, as B-mode images are log-compressed, which alters the statistical relationship between pixels. Therefore, there is a need for a method that can generate Nakagami images from log-compressed B-mode images in real-time, thereby accelerating the implementation of Nakagami images in clinical ultrasound systems by demonstrating their utility in various diseases using the vast amount of available B-mode image data in clinics.

In addition to ultrasound-based multi-parametric approaches, tumor shape features have been shown to enhance diagnostic accuracy, as evidenced by studies that link morphological characteristics to malignancy. Irregular shape and spiculated margins are strongly associated with malignant tumors. One study found that Fourier-based shape analysis improved classification accuracy by 17.32% compared to traditional shape metrics, such as the lobulation index, in distinguishing between malignant and benign lesions [14]. Furthermore, integrating shape features (e.g., oval vs. irregular) with BI-RADS classification reduced false-positive biopsies by 27.8% in AI-assisted systems, demonstrating their complementary role in refining diagnostic precision [15]. Classical region, threshold, and edge-based techniques have been applied to segment breast tumors on MRI, ultrasound, and mammography images [16]. Classical threshold- and edge-based techniques (e.g., region-growing, active contours) are sensitive to initialization and artifacts, often achieving moderate Dice scores (50–75%) [17], [18]. While deep learning models, such as U-Net, improve performance (Dice: 0.77–0.89) [19], [20], they still struggle with irregular morphologies and shadowing artifacts, resulting in false positives in 8–27% of cases [21], [22]. Recent advances in breast ultrasound tumor segmentation utilize hybrid deep learning architectures to address challenges such as speckle noise and ambiguous boundaries. Various deep learning-based methods have been developed to improve breast tumor segmentation in ultrasound images. These include fuzzy logic-enhanced CNNs [23], Res Path and dense block-integrated U-Nets [24], small tumor-focused networks like ESTAN [25], and attention-augmented models such as Sharp Attention U-Net [26], achieving Dice scores ranging from 0.81 to 0.94. While these approaches have improved robustness over classical methods (Dice: 0.50–0.75), segmentation can be further enhanced by employing a streamlined encoder-decoder architecture that enables multiscale feature extraction and maintains consistency between encoding and decoding paths. This design minimizes semantic gaps and improves accuracy across diverse tumor morphologies, without relying on complex attention mechanisms or fuzzy logic modules [27]. Instead of using B-mode, QUS, or tumor shape parameters separately, combined methods that integrate U-Net segmentation with Nakagami parametric imaging can improve classification accuracy. This study presents a deep learning-based framework to discriminate breast masses by integrating B-mode and Nakagami imaging with automatic lesion segmentation. Nakagami imaging was chosen because it provides quantitative information regarding tissue microstructure, while lesion shape provides critical distinguishing features between benign and malignant masses. We developed three deep-learning models: NakaSynthNet, SegmentNet, and FeatureNet. NakaSynthNet generates pseudo-Nakagami images from the log-compressed B-mode images in real time. True Nakagami images are generated from the beam-formed envelope-detected RF image (i.e. B-mode without log-compression). Therefore, NakaSynthNet will enable the use of massive B-mode image data that is available in clinics without requiring RF data. The SegmentNet model is a typical U-Net design with post-processing capabilities, enabling the segmentation of multiple lesions from a B-mode image. The FeatureNet model employs a transfer learning approach, merging the two features —Nakagami generation and segmentation—to classify breast masses.

These models were trained on both simulated and clinical ultrasound images. To address the data scarcity, a large dataset was generated by converting natural fruit images and mammography images into simulated ultrasound images using Field II [28], [29]. Echo signal-to-noise ratio (SNR) and intensity of target regions (i.e., fruits and tumors) varied to create a diverse dataset.

In summary, this study presents a comprehensive framework for discriminating breast masses using multi-parametric images, lesion shape, and deep learning, addressing the gaps in data variability. Due to the robustness of this model, some of the methods developed in this study can be used in other disease applications.

## II. METHODOLOGY

### A. Overall Architecture

Fig. 1 illustrates the simplified overview of the proposed deep learning framework, which incorporates three distinct CNN architectures: NakaSynthNet, SegmentNet, and FeatureNet. NakaSynthNet was trained to learn the mapping of Nakagami parameter images from conventional B-mode ultrasound images. True Nakagami was calculated from the envelope-detected RF data without log compression, which will be denoted as uBmode (i.e., uncompressed Bmode) throughout the manuscript. Note that B-mode implies that log compression is applied to the enveloped detected RF data. SegmentNet was trained to perform automated lesion segmentation using paired B-mode images and corresponding manually annotated lesion masks. FeatureNet extracts and integrates both quantitative features from the predicted Nakagami parameter images and qualitative features from the segmented lesion masks. These composite features are then used as input to a classification network employing transfer learning with a pre-trained VGG-16 model. The dataset consists of in vivo clinical B-mode images and Field II [28], [29] simulated B-mode images of natural fruits, breast masses from mammography images, and digital phantoms containing lesions.

**Fig. 1.**
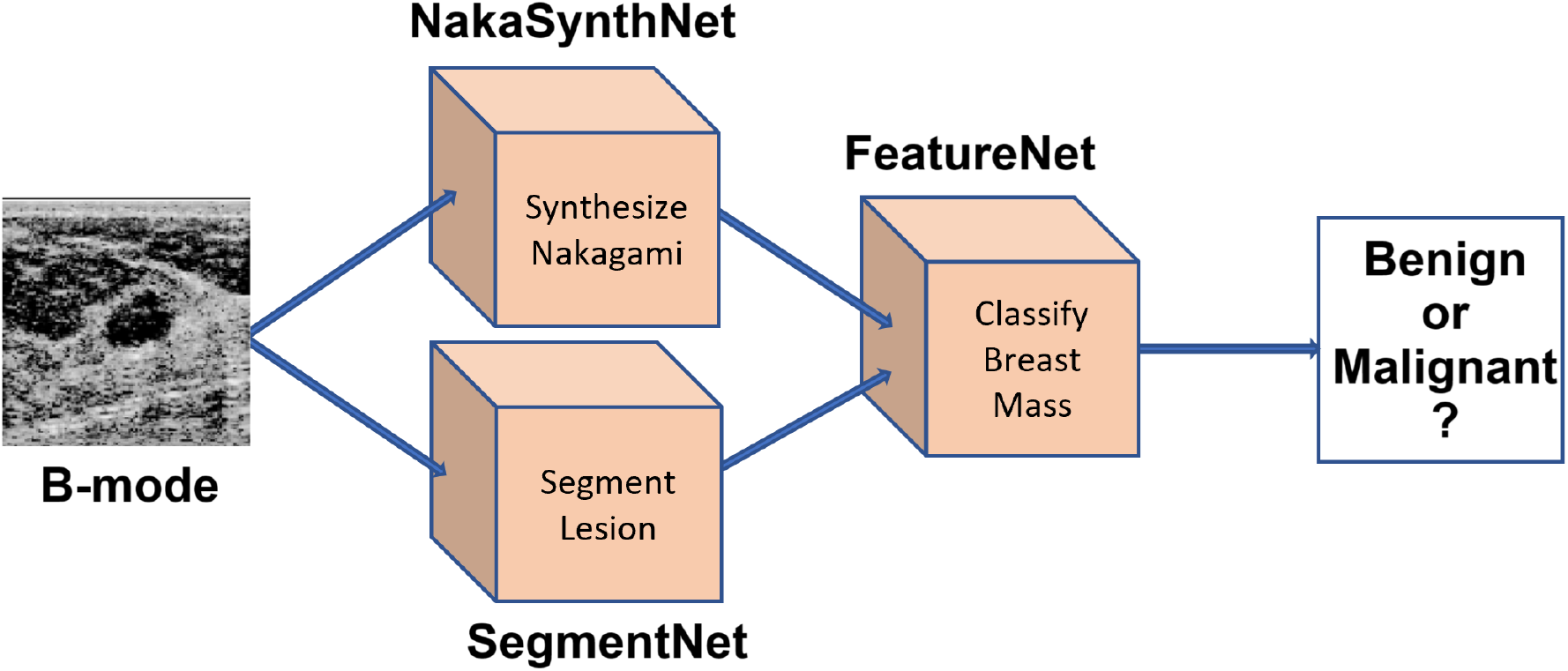
Overall framework of the proposed deep learning framework.

#### 1) NakaSynthNet

The key components of the NakaSynthNet architecture are Nakagami conversion, superimpose layer, convolutional layer, dense layer, and a composite loss monitoring function (Fig. 2a). The true Nakagami image was generated by fitting the pixel intensities of a square window of a uB-mode image to the Nakagami statistical model and then sliding the window over the whole uB-mode image [13], [30]. During training, paired Nakagami and B-mode images were weighted and superimposed via superimpose layers to retain the structural detail inherent in the B-mode while incorporating quantitative tissue information from the Nakagami map. The architecture was trained end-to-end using convolutional layers and used the final dense layer to predict the Nakagami image corresponding to the input B-mode image. During inference, the network accepts a B-mode with unknown log compression level and generates a Nakagami parameter map without requiring access to the original uB-mode data. The architecture adopts a dual-branch design that combines prediction and fusion. The prediction branch uses an encoder–decoder structure to generate a Nakagami-like statistical map from the input B-mode image. It employs 3×3 convolutional layers with increasing filter depths (32, 64, 128) to extract hierarchical features, followed by transposed convolutions for up-sampling. A final 3×3 convolution with sigmoid activation outputs a three-channel RGB Nakagami image normalized to the [0, 1] range. To support both training and inference modes, a switching mechanism checks the maximum intensity in the externally provided Nakagami map (i.e., during training). If the input map contains only zeros (i.e., during testing), the model defaults to its own predicted map. A trainable fusion layer then combines the B-mode image with the selected Nakagami map using learnable scalar weights, balancing anatomical detail with statistical texture. The fused output is passed through additional convolutional layers (128, 64, 32, and 16) to produce the final result: a colorized Nakagami “super-image” that integrates both structural and statistical information. The model is trained using a composite loss function that combines mean squared error (MSE) and structural similarity index (SSIM) to guide the prediction of both the Nakagami map and the RGB output image. This dual-loss approach encourages the network to maintain both pixel-level accuracy and perceptual similarity to the ground truth. Training is performed using the Adam optimizer with a learning rate of 0.001, along with early stopping and learning rate reduction to prevent overfitting and enhance generalization. To evaluate robustness, the model was trained and tested using multiple Nakagami window sizes (2, 3, 4, and 5). During training, the model simultaneously learned to predict the Nakagami parameters, their corresponding RGB representations, the optimal window size for Nakagami estimation, and the fusion weights for combining B-mode and Nakagami images. Final performance metrics, including accuracy, SSIM, and MSE, were obtained by averaging results over five independent runs. We also trained the model using uB-mode to predict Nakagami, evaluating the performance loss of models with uB-mode versus B-mode input. This model with uB-mode will be used if the RF data is available, allowing for the real-time generation of the Nakagami image.

**Fig. 2.**
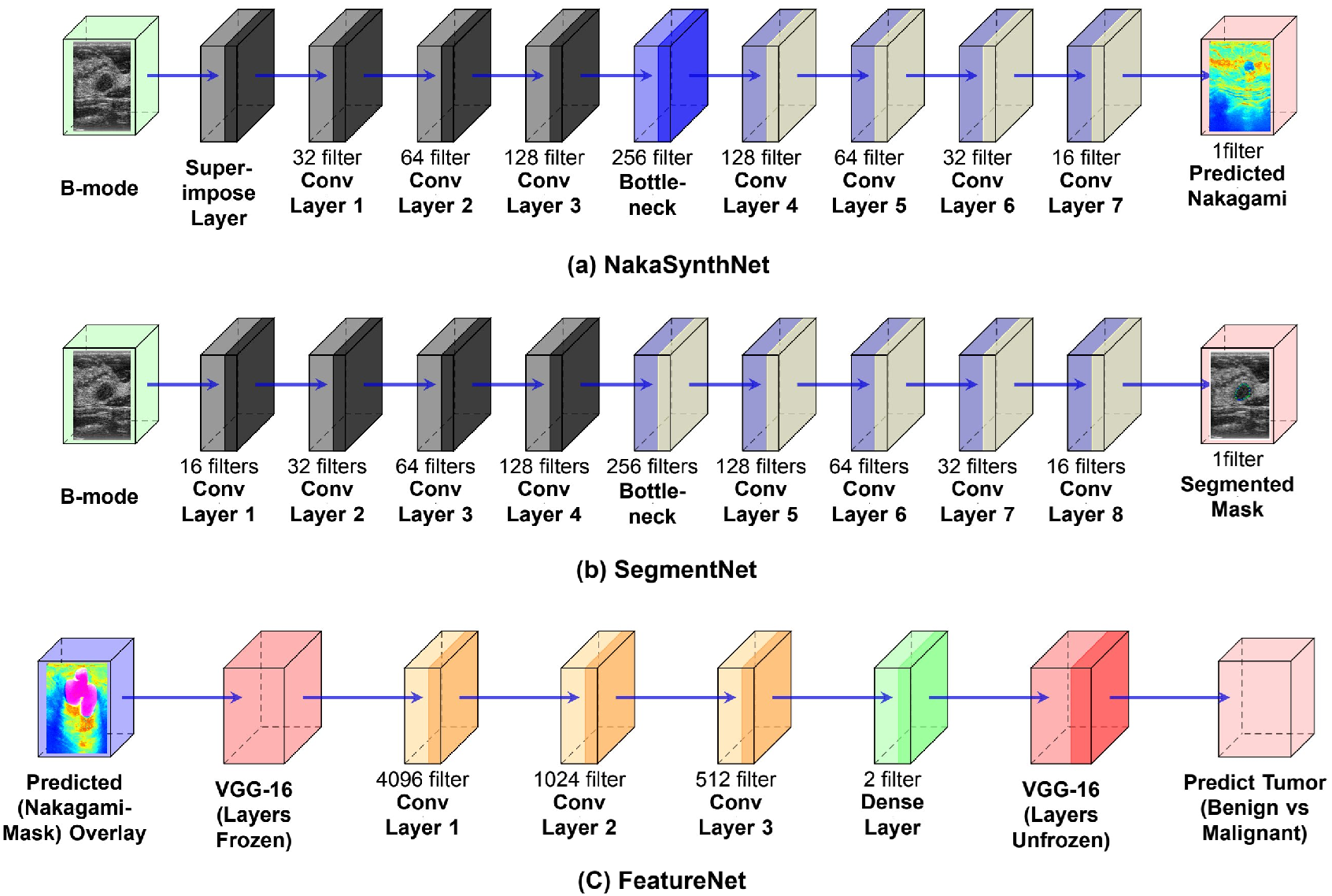
Complete Model Architecture of (a) NakaSynthNet, (b) SegmentNet (Encoder: Layer 1-4, bottleneck, Decoder: Layer 5-8), and (c) FeatureNet with VGG-16.

#### 2) SegmentNet

SegmentNet (Fig. 2(b)), a custom CNN based on the attention U-Net design [43], is trained using paired B-mode images and manually annotated lesion masks, enabling it to predict segmentation masks from B-mode inputs during inference. The network features an encoder–decoder structure with skip connections, allowing a balance of feature extraction and spatial detail recovery. The encoder utilizes convolutional blocks with 3×3 kernels and progressively increasing filter sizes (16, 32, 64, 128), each followed by batch normalization, ReLU activation, and 2×2 max pooling for down-sampling. A dropout rate of 0.1 is applied after each pooling step to reduce overfitting. This hierarchical feature effectively extracts both low-level details, such as edges, and high-level abstract information, such as object shapes. At the center of the network, a bottleneck layer with 256 filters captures high-level semantic features, serving as a compressed global representation of the input before spatial details are recovered in the decoder. It filters out irrelevant information and enhances critical segmentation cues for accurate localization of lesion boundaries. The decoder then progressively restores spatial resolution using transposed convolutions (3 × 3 kernel, stride 2), with decreasing filter sizes (128, 64, 32, 16). Each decoder block concatenates its upsampled features with outputs from the corresponding encoder layer via skip connections, allowing for the recovery of fine structures, such as lesion boundaries. The final output is generated through a 1 × 1 convolution with sigmoid activation, producing a binary mask where white regions (value 1) indicate lesions and black regions (value 0) indicate background. The model is trained using binary cross-entropy (BCE) loss, optimized with the Adam algorithm (learning rate 0.0001), on input images of size 256 × 256 × 1. Training is performed for up to 50 epochs with a batch size of 16 and early stopping (patience = 10) to prevent overfitting. This architecture effectively balances hierarchical feature extraction with spatial reconstruction, making it well-suited for accurate and efficient segmentation of complex anatomical structures in medical ultrasound images. Similar to the NakaSynthNet, SegmentNet was also trained with uB-mode separately to evaluate the difference in performance when B-mode versus uB-mode was used in training.

#### 3) FeatureNet

The FeatureNet model (Fig. 2 (c)) classifies breast masses as benign or malignant by integrating information from two complementary inputs: NakaSynthNet-predicted Nakagami images and SegmentNet-derived lesion masks. These are superimposed to create a composite RGB image that combines quantitative tissue characterization with tumor spatial location and morphological characteristics. These fusion highlights diagnostically relevant regions by preserving structural boundaries while incorporating backscatter texture. These overlaid images were input to the FeatureNet, which includes a VGG-16, a well-established CNN architecture known for its deep feature extraction capabilities, pretrained on the ImageNet dataset [31].

The composite image is resized to 224 × 224 and fed into a VGG-16-based CNN pretrained on ImageNet. Initially, the 13 convolutional layers and 5 max-pooling layers of VGG-16 are frozen to retain general visual features by reducing spatial dimensions while preserving important spatial hierarchies. The extracted deep features are passed through dense layers of 4096, 1024, and 512 units, each followed by ReLU activation and dropout (rates: 0.3, 0.2, 0.1) to prevent overfitting. After this initial phase, VGG-16 layers are unfrozen for fine-tuning, allowing domain-specific adaptation. A final dense layer with softmax activation outputs class probabilities for benign and malignant categories.

The model is trained using sparse categorical cross-entropy loss and the Adam optimizer, with a learning rate of 0.0001 during pretraining and 0.00001 during fine-tuning, over 50 epochs each. Data augmentation techniques, such as rotation, zooming, flipping, and shifting, were applied to enhance generalization and robustness. The use of dropout and two-phase training, with and without frozen layers, helps prevent overfitting, especially when training on relatively small medical image datasets. The large initial dense layer captures global image patterns, while subsequent layers refine features critical for lesion classification. The combination of transfer learning, feature fusion, and regularization allows FeatureNet to generalize well even on limited medical imaging datasets. Performance is evaluated using accuracy, sensitivity, specificity, and the area under the curve (AUC), ensuring a reliable assessment of clinical relevance.

FeatureNet was trained separately on eight combinations of Nakagami, B-mode, and lesion masks to identify the performance of the proposed lesion masks overlaid on Nakagami images compared to other combinations. However, the number of images varied between this combination as the RF data was not available in the 1st clinical ultrasound dataset (sec section B.1). Here are the combinations: (i) Nakagami synthesized from B-mode + lesion masks (NBLM, N = 747); (ii) B-mode + lesion masks (BLM, N = 747 images); (iii) NBLM (N = 100); (iv) Nakagami synthesized from uB-mode + lesion masks (NuBLM, N = 100); (v) B-mode + lesion masks (BLM, N = 100 images); (vi) uB-mode + lesion masks (uBLM, N = 100 images); (vii) B-mode (N = 100); (viii) B-mode (N = 747). Combinations (iii) and (v) were subcategorized based on log compression levels: −50 dB, −60 dB, −70 dB, and −80 dB. Apart from these, all the combinations that contain B-mode, have the log compression levels of: −50 dB, −60 dB, −70 dB, and −80 dB for the 2nd clinical ultrasound dataset of 100 images (section B.1), as RF data were available only for this dataset. Note that the combination numbers (i) and (iii) are the same in terms of input, with only a difference in the number of images, 100 versus 747. We retrained and retested the combination number (i) using 100 images to ensure a fair comparison with combination number (iv) as the RF data only available for 100 images. The combination number (viii) was used to assess the accuracy of multimodal imaging with lesion masks compared to B-mode only. The dataset was split into 75% for training and 25% for testing.

### B. Dataset

We used a total of 110,247 images from four sources: (1) publicly available clinical in-vivo B-mode images of breast masses and synthetic B-mode images of (2) natural fruit image, (3) breast masses derived from clinical mammogram images, and (4) digital phantoms containing lesions generated using Field II [28], [29] with L7-4 transducers (Philips Healthcare, Andover, MA, USA).

#### 1) Clinical B-mode images

We used two clinical in vivo breast ultrasound datasets [32], [33]. The BUSI dataset [32], contains 780 ultrasound images from 600 women aged 25–75 years, collected in 2018. We selected 647 images, labeled as either benign or malignant, for this study. Normal breast images from the dataset were not used. As the RF data was not available for this dataset, we used it only for the SegmentNet and FeatureNet. The second dataset [33] comprises 100 ultrasound RF data sets from 78 women, collected between 2013 and 2015, with 52 malignant and 48 benign cases. RF data were acquired using an L14-5/38 linear array transducer. B-mode images were reconstructed using the Hilbert transform, followed by log compression at −50, −60, −70, and −80 dB. Thus, total number of clinical images in the dataset was 647 + 100 * 4 = 1047. Nakagami was calculated using both uB-mode and B-mode data. This dataset is used in all 3 models.

#### 2) Ultrasonic simulation of natural fruit images

B-mode images of 200 natural fruit images [34] were simulated using the Field II with an L7-4 linear array transducer (128 elements, 5.2 MHz center frequency, and 100 MHz sampling frequency) (Fig. 3), similar to the method described in [35].

**Fig. 3.**
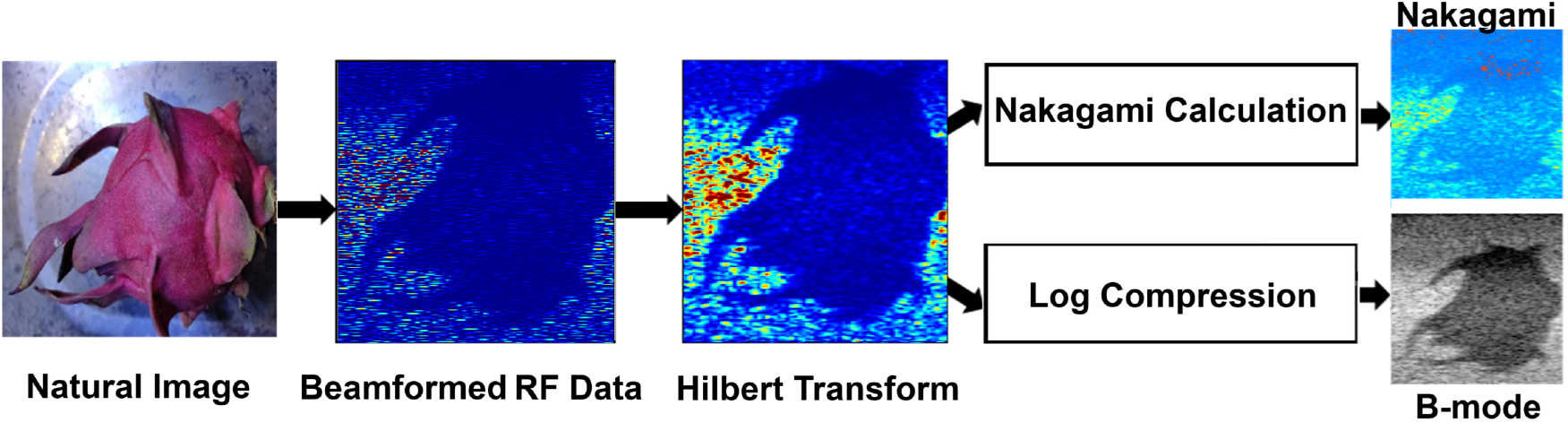
Steps for generating B-mode and Nakagami images from the natural images in Field II.

The fruit image is converted to a binary image to treat the fruit region and background as a lesion and the tissue surrounding the lesion, respectively. A total of 68,475 scatterers (i.e., reflecting 15 scatterers/resolution cell) with randomly varying amplitudes were placed randomly in a 20 mm × 30 mm (axial × lateral) area by scaling the fruit image to that area. The mean lesion scatterers’ intensity was varied from −30 to 30 dB in 10 dB increments, except 0 dB, with respect to the background. Therefore, the lesion will be hypoechoic when the lesion’s scatterers’ intensity is negative dB with respect to the background and vice versa.

A Hanning-windowed sinusoidal pulse was used for excitation, with dynamic focusing and apodization to enhance lateral resolution. Sixty-seven RF lines with an F-number of 1.5 and a focal depth of 20 mm were simulated via parallel processing (using the parfor function in MATLAB) by translating the focus across the lateral field of view to generate a single image. Additive white Gaussian noise (awgn function) was added to the beamformed RF data at SNR levels ranging from 5 to 35 dB in increments of 5 dB. The absolute Hilbert transform was applied to the beamformed RF data to obtain envelope data (i.e., uB-mode). Both uB-mode and uB-mode with log compression levels of −50 dB, −60 dB, −70 dB, and −80 dB were saved. Nakagami parameters were calculated from the uB-mode. Therefore, 168 B-mode images (6 scatterer intensities, 7 SNRs, and 4 log compression levels) were generated from a single fruit image, resulting in a simulated dataset of 33,600 images (200 × 168) to make the dataset robust and diverse for NakaSynthNet and SegmentNet models.

#### 3) Ultrasonic Simulation of Mammogram-Derived Breast Mass Images

A total of 200 breast mass images were selected from a larger mammogram-derived dataset containing 10,239 images [36]. These images were simulated using the same Field II setup and methodology described for the natural fruit dataset (L7-4 transducer, varying scatterer intensity from −30 to +30 dB, SNR from 5 to 35 dB, and log compression at −50, −60, −70, and −80 dB). This process yielded 33,600 B-mode images (200 × 168), which were used to increase diversity in training the NakaSynthNet and SegmentNet.

#### 4) Ultrasonic simulation of Digital lesion phantoms

Using previously described methods, ultrasound simulation of 250 lesion phantoms was performed using Field II with an L7-4 transducer. Lesion shape (e.g., circular with smooth or rough edges) and size (e.g., diameters varied from 6 mm to 12 mm) were varied to have a robust and diverse dataset. Similarly, the intensity, SNR, and Log compression labels were varied to generate 42,000 simulated B-mode images (250 × 168) used in SegmentNet and NakaSynthNet.

## III. RESULTS

### 1) NakaSynthNet

Fig. 4 illustrates the effectiveness of the NakaSynthNet model in synthesizing Nakagami images from B-mode ultrasound images of benign breast mass, regardless of the log compression level in B-mode. Three observations are notable. First, predicted Nakagami from −50 dB B-mode closely resembles the true Nakagami compared to other log compression labels. Note that true Nakagami was calculated from the uB-mode. Second, while predicted Nakagami intensities were generally highest from −70 dB B-mode compared to the other log compression levels, lesions are still discernible in all predicted images. The (MSE and SSIM) of the predicted Nakagami values at −50, −60, and −70-dB B-mode were (3% and 96%), (2% and 97.9%), and (1% and 98.6%), respectively. Third, the MSE between predicted Nakagami at −50 dB versus −60 dB, −50 dB versus −70 dB, and −60 dB versus −70 dB was 1.5%, 3%, and 2%, respectively. Fourth, histogram plots indicate that 1.5% of pixels exhibit a 10% difference in intensity between the predicted and true Nakagami values, irrespective of the log compression level.

**Fig. 4.**
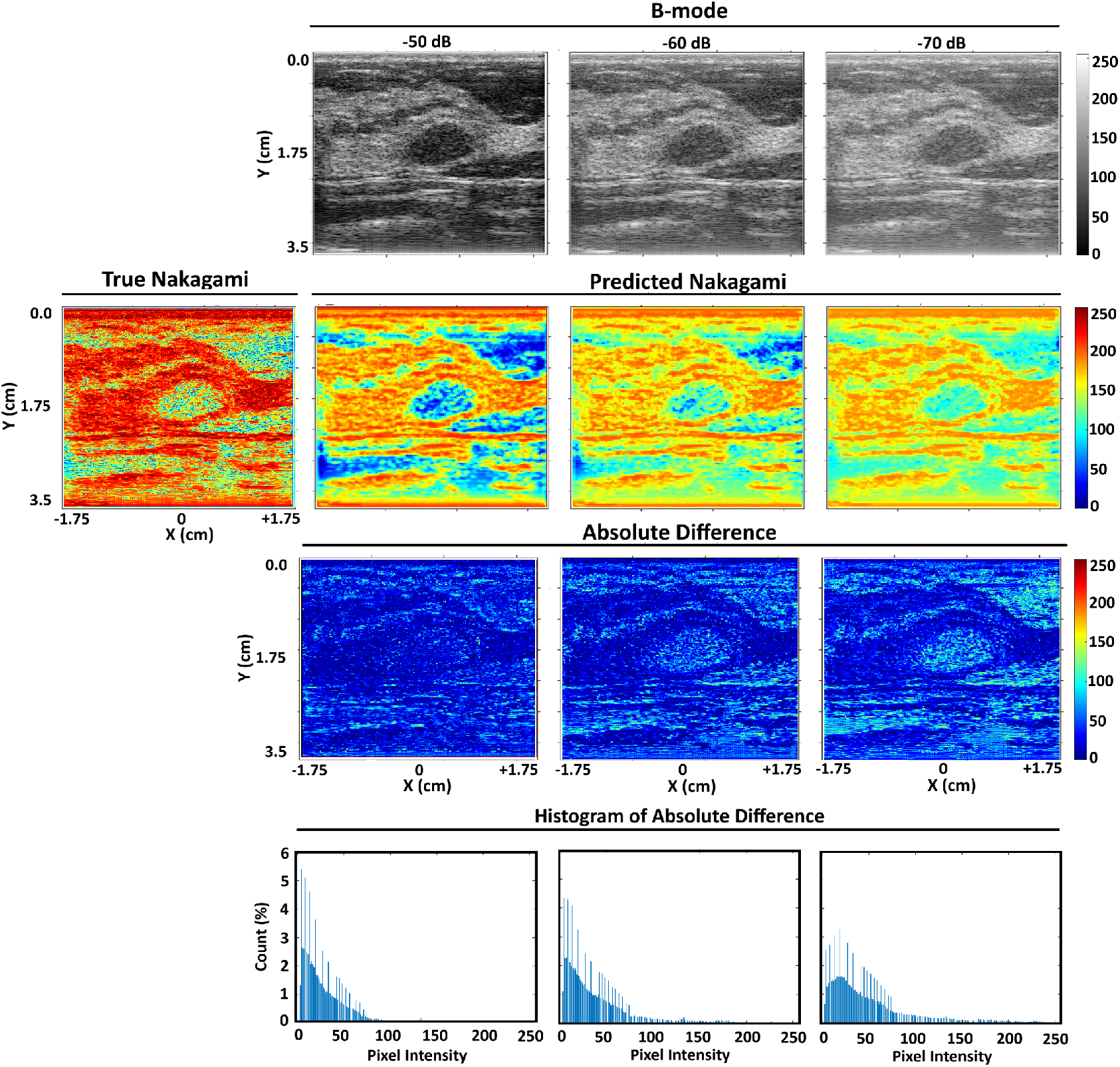
Representative B-mode (1st row), true Nakagami and predicted Nakagami (2nd row), and absolute difference between true and predicted Nakagami (3rd row) images, and histogram (4th row) of the absolute difference images of benign breast mass with log-compression of −50 dB, −60 dB, and −70 dB.

Overall, in the test set, NakaSynthNet demonstrated an average SSIM of 98% and MSE 0.09%, affirming the model’s high performance and robust capability to produce reliable Nakagami images from B-mode images when RF data is not available. However, when the RF data was available, i.e., NakaSynthNet predicted Nakagami from the uB-mode, the performance improved, as indicated by the SSIM and MSE of 99.3% and 0.07%. The NakaSynthNet needs, on average, 46 ms (i.e., 21 frames per second) to generate a Nakagami image compared to 90 ms (i.e., 11 frames per second) when the Nakagami equation [41] is used to generate the image. The computation is performed on an AMD Ryzen Threadripper PRO 7965WX 4.20 GHz processor with 24-Cores cores, 144 GB of RAM, and 8 GB GPU configured computer. Therefore, NakaSynthnet allows the generation of images in real-time.

### 2) SegmentNet

Fig. 5 qualitatively demonstrates that SegmentNet accurately predicted the location, shape, and size of benign and malignant lesions compared with the true lesion masks provided in the dataset. The SegmentNet’s (accuracy and pixel-wise binary cross-entropy loss) were (98.8% and 1.2%), (95.2% and 4%), (99.8% and 0.1%), and (97.6% and 3%) for these four representative images.

**Fig. 5.**
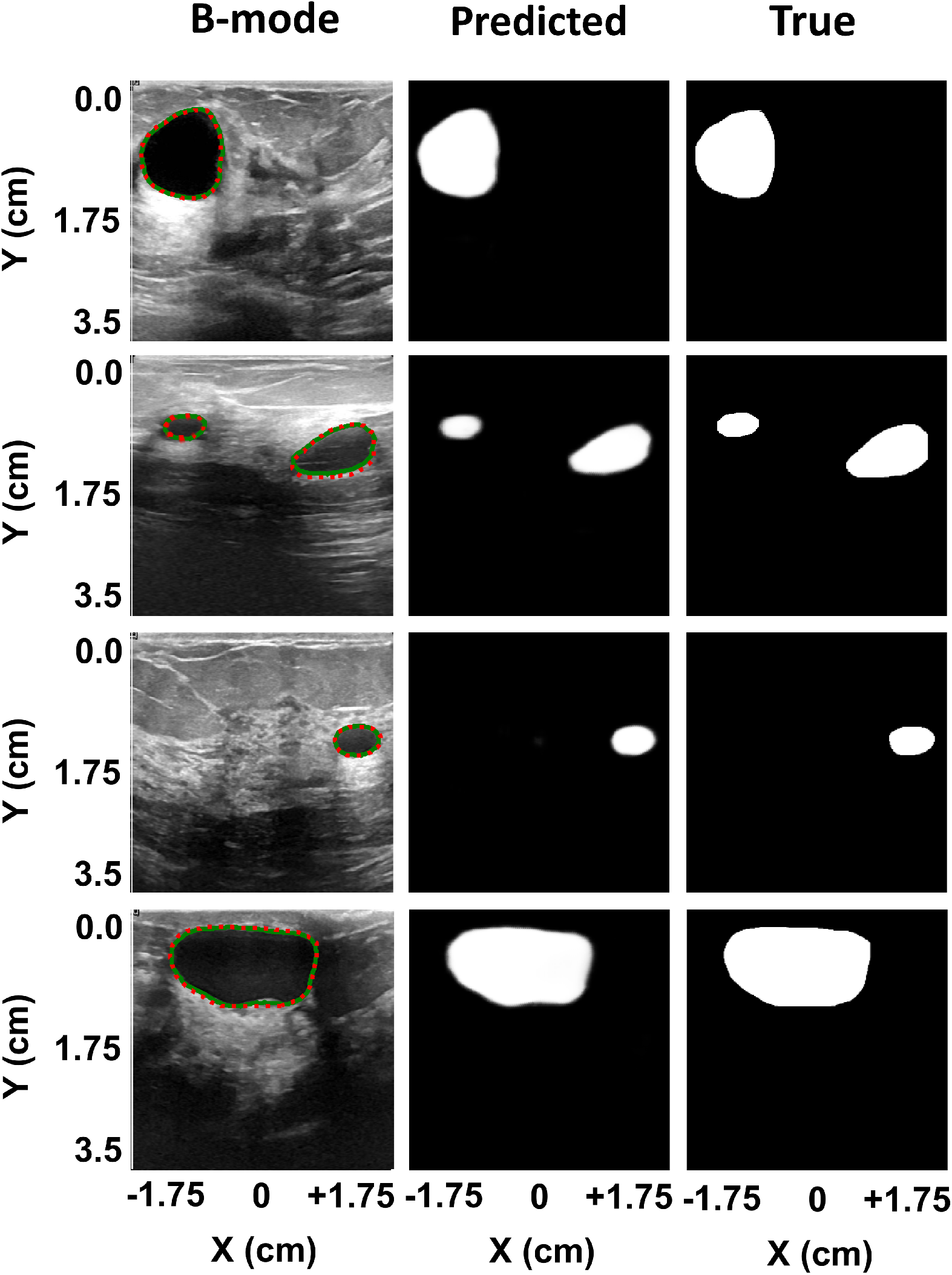
Representative B-mode of benign (row 1-3) and malignant (row 4) with true (red) and predicted (green) tumor boundaries (left column), predicted mask (center column), and true mask (right column).

Overall, in the test set, SegmentNet achieved an average accuracy of 98.4% and pixel-wise binary cross-entropy loss of 0.4%, with a dice similarity coefficient of 94%, demonstrating its accuracy in delineating lesion boundaries. However, when the uB-mode data was used as input, the performance degraded, as indicated by the accuracy and binary cross-entropy loss of 96.7% and 10%, respectively (see Fig. 7). The SegmentNet took on average 50 ms (i.e., 20 frame/s) to generate the lesion masks on the same computer.

### 3) FeatureNet

Table I provides a comprehensive performance comparison of FeatureNet using various input combinations and preprocessing schemes. When utilizing Nakagami from B-mode with Lesion Mask (NBLM), the network achieved the highest performance across all evaluation parameters: an accuracy of 96.7%, sensitivity of 93%, specificity of 98%, and AUC of 98% on the largest dataset (N = 747). Furthermore, NBLM exhibited remarkable robustness; its performance remained nearly unchanged when the data size was reduced from over 747 to 100 (accuracy: 96.5% vs. 95%). In contrast, BLM (B-mode with Lesion Mask) performance was consistently lower than NBLM across all metrics but superior to B-mode alone and to inputs relying on uncompressed B-mode data (NuBLM/uBLM). Stratification by log-compression level within N = 100 samples showed that NBLM retained stable accuracy (±1–2%) for lower log compression (−50 dB, −60 dB) ranges, while BLM experienced accuracy degradation (down to 73% at –80 dB). Notably, models using NuBLM and uBLM inputs—derived from uncompressed B-mode—consistently underperformed, both in sensitivity (70% and 67%), specificity (75% and 89%) and AUC (68% and 70%), relative even to B-mode alone. B-mode alone maintained better performance than NuBLM or uBLM with accuracy values of 90%, confirming the advantage of conventional log-compression. Cross-validation showed that NBLM’s predictions were more consistent, compared to higher variability in BLM and NuBLM/uBLM. On the test set of 150 cases, FeatureNet with NBLM correctly classified 97% of cases (145/150), misclassifying only 2 malignant and 3 benign cases. The processing time averaged 56 ms per case.

**TABLE I.** ERFORMANCE COMPARISON OF FEATURENET FOR DIFFERENT COMBINATIONS (NBLM = Nakagami FROM B-mode WITH Lesion Mask, BLM = B-mode WITH Lesion Mask, NuBLM = Nakagami FROM uB-mode WITH Lesion Mask, uBLM= uB-mode WITH Lesion Mask)

| Evaluation Parameters | NBLM | BLM | NBLM |  |  |  |  | BLM |  |  |  |  | B-mode |  | NuBLM | uBLM |
| --- | --- | --- | --- | --- | --- | --- | --- | --- | --- | --- | --- | --- | --- | --- | --- | --- |
|  |  |  | -50 dB | -60 dB | -70 dB | -80 dB | Random | -50 dB | -60 dB | -70 dB | -80 dB | Random | Set 1 | Set 2 |  |  |
| Data (N) | 647+<br>100 | 647+<br>100 | 100 | 100 | 100 | 100 | 100 | 100 | 100 | 100 | 100 | 100 | 647+<br>100 | 100 | 100 | 100 |
| Accuracy (%) | 96.7 | 86 | 95 | 90 | 85 | 75 | 95 | 85 | 83 | 80 | 73 | 83 | 90 | 90 | 90 | 90 |
| Sensitivity (%) | 93 | 85 | 88 | 85 | 82 | 78 | 85 | 83 | 82 | 80 | 76 | 82 | 76 | 77 | 70 | 67 |
| Specificity (%) | 98 | 80 | 99 | 99 | 95 | 92 | 98 | 80 | 82 | 81 | 78 | 80 | 90 | 90 | 75 | 89 |
| AUC (%) | 98 | 90 | 98 | 94 | 88 | 80 | 98 | 90 | 88 | 80 | 80 | 90 | 70 | 72 | 68 | 70 |

In summary, these observations collectively demonstrate that Nakagami synthesized from log-compressed B-mode + lesion mask (NBLM) yields the highest, most stable diagnostic accuracy, outperforming all other input combinations. For the best-performing input (i.e., NBLM), the training accuracy remained higher than the validation accuracy after epoch #11, indicating that the model was not overfitted (Fig. 6 (a)). At some epoch, the model slightly overfitted but that was due to the dataset imbalance (benign dataset was almost 2x larger than malignant dataset). Compared to the imbalance, the model overfitted very slightly at those epochs. Out of 150 test cases, FeatureNet with NBLM inputs identified 145 cases (i.e., 97%) correctly, 2 malignant cases as benign, and 3 benign cases malignant (Fig. 6 (b)). The FeatureNet took, on average, 56 ms (i.e., 18 frames/s) to predict cases on the same computer.

**Fig. 6.**
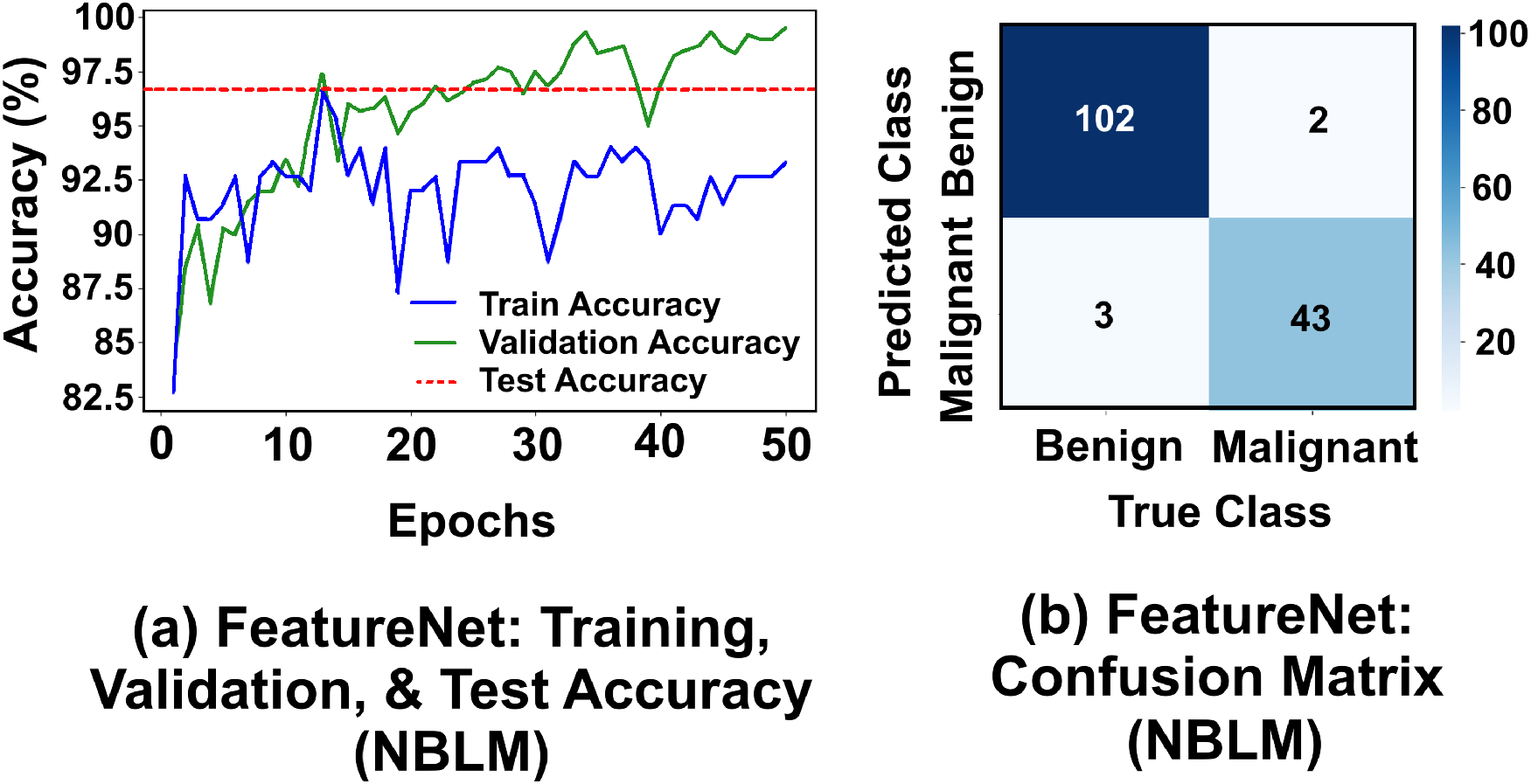
(a) Training, validation, and test accuracy of FeatureNet versus epochs (b) Confusion matrix for the FeatureNet.

**Fig. 7.**
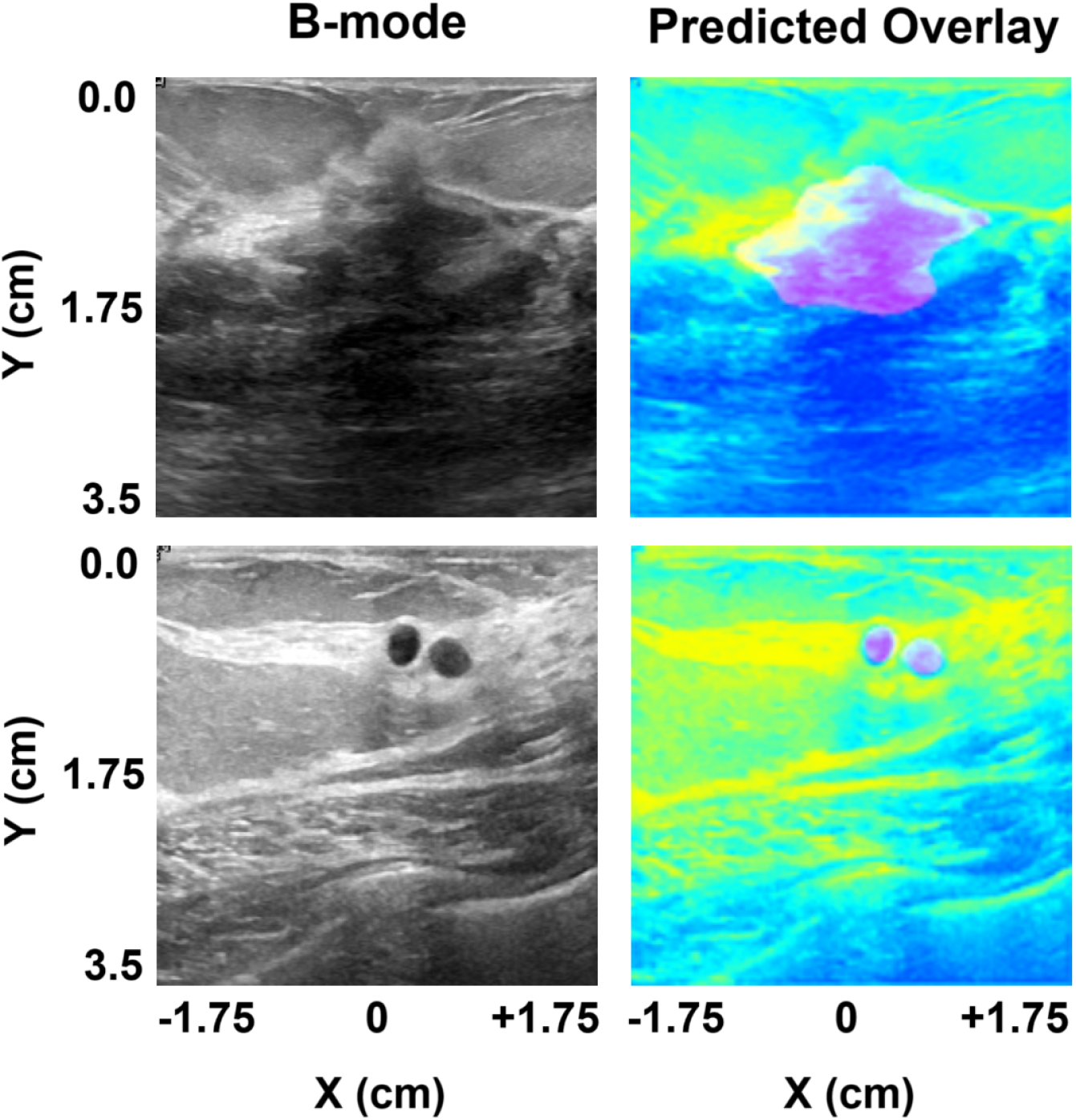
Output of the proposed framework for benign and malignant cases after combining all three networks. The frameworks demonstrate a lesion mask overlaid on the Nakagami image and prediction when the input is B-mode.

### 4) Combined Output of All Three Models

Fig. 7 illustrates how the three models can be combined to make a prediction when the input is B-mode. NakaSynthNet and SegmentNet will generate the Nakagami and lesion mask, respectively. A computer algorithm will then overlay the lesion mask over the Nakagami for input to FeatureNet. The frameworks will demonstrate both the overlay image and FeatureNet’s prediction. It took, on average, 56 ms (i.e., 18 frames/s) to generate the overlay image and make a prediction from the input B-mode image.

## IV. DISCUSSION

We proposed a deep learning-based framework for breast cancer diagnosis by incorporating morphological characteristics of breast lesions and quantitative ultrasound backscatter information in terms of Nakagami parameters. To the best of our knowledge, this is the first study to combine lesion mask and Nakagami parameters by predicting them solely from the B-mode images, without access to raw RF data, rather than manually annotating or calculating them. Previous deep learning studies have either used B-mode alone [5], [37], Nakagami alone [11], [13], [30], lesion mask alone [5], or a combination of B-mode and lesion masks [5], [17], [38]. For the segmentation of tumor, different custom U-Net, attention U-Net, ResNet, Fuzzy models, etc are used [39] [40], and for the diagnosis, LeNet, different regression models, SVM, k-NN, ANN, CNN models, transfer learning models like VGG-16, VGG-19 etc are widely used [41]. Note that FeatureNet performed the main diagnostic decision in our proposed framework. This model also features a custom CNN network based on VGG-16 transfer learning. However, this model combines the performances of NakaSynthNet and SegmentNet, which predict the Nakagami image and lesion mask respectively, from the B-mode. The prediction of Nakagami and lesion masks provides real-time imaging and segmentation of lesions.

As the datasets differ between the proposed and previous studies, it is difficult to compare their performance directly. That’s why we compared the performance of FeatureNet with different input combinations. The results in the table highlight the superiority of using NBLM inputs—Nakagami features synthesized from log-compressed B-mode images with lesion masking—over all other tested configurations for breast cancer diagnosis with FeatureNet. Firstly, these results demonstrate that quantitative Nakagami features derived from log-compressed B-mode images offer both exceptional accuracy and remarkable robustness to dataset size reductions, log-compression dynamic range changes, and train-test splits. Secondly, though traditional approaches use uncompressed B-mode data to compute Nakagami features (NuBLM/uBLM), our findings show that such approaches produce inferior diagnostic performance in our dataset. This is likely because uncompressed data is susceptible to noise and dynamic-range artifacts, resulting in poor stability and lower AUC values. Remarkably, even using only the B-mode image input outperforms the NuBLM/uBLM configurations, reaffirming the clinical advantage of log-compression preprocessing. Third, the fusion of qualitative (image structure) and quantitative (Nakagami statistics) features—achieved by NBLM—proves more effective than qualitative-only (B-mode), or quantitative-only (Nakagami from uB-mode) inputs. The difference becomes more pronounced for small or low-contrast lesions. This suggests that Nakagami-based features are particularly effective in settings with low structural visibility. Fourth, the model trained with NBLM exhibited substantially lower variance across random splits, indicating much more stable and predictable learning. Although generating NBLM features requires additional computation (roughly double that of BLM), this is more than justified by the substantial gains in performance and robustness—particularly when compared to the even greater computational burden of NuBLM/uBLM, which do not confer any performance benefit. Lastly, while a slight overfitting was observed as the training accuracy occasionally exceeded validation accuracy, this was attributed primarily to class imbalance rather than architectural inadequacy. The model’s generalizability remains strong, with minimal overfitting, as evidenced by its test set performance. In summary, NBLM is the optimal FeatureNet input for breast cancer diagnosis, consistently yielding the highest accuracy, stability, and robustness among all feature combinations in our dataset.

While the FeatureNet performs better with NBLM, our proposed NakaSynthNet can synthesize Nakagami images from either B-mode or uB-mode with similar performance with similar performance (e.g., SSIM of 98% and 99.3% for B-mode and uB-mode) in real-time (Fig. 4), making NakaSynthNet a universal tool for other clinical applications. Nakagami parameters have been demonstrated to improve performance in fibrotic liver tissue [42], benign and malignant breast tumors [13], [30], kidney stone detection [43], abdominal imaging [44], and diagnostics for Duchenne muscular dystrophy [45] and thyroid nodules [46]. diseases. NakaSynthNet will enable researchers to demonstrate the relevance of Nakagami parameters in diagnostics by leveraging the vast B-mode data available in clinics. NakaSynthNet can be easily integrated into any ultrasound system without modifying its ultrasound data processing pipeline. NakaSynthNet can be added after generating the B-mode image to synthesize the Nakagami image.

Synthesis of one medical imaging modality to another (e.g., CT to MRI) [47] or B-mode generated from research to clinical ultrasound systems [48] has been demonstrated previously [49]. Unlike prior works—such as CycleGAN-based imaging translation model [47] for cross-modality mapping between CT and MRI, GAN framework [48] for synthesizing realistic ultrasound images and segmentation labels to address annotation scarcity, and MimickNet [49], which emulates clinical ultrasound post-processing pipelines through black-box deep networks—the proposed NakaSynthNet is purpose-built as a dual-branch encoder–decoder CNN with a dedicated fusion layer to directly synthesize quantitative Nakagami parameter maps from log-compressed B-mode images, requiring neither raw RF data nor adversarial training. Rather than generating visually plausible images or labels, NakaSynthNet uniquely integrates anatomical and statistical information via learnable superimpose layers, enabling real-time, diagnostically meaningful quantitative ultrasound imaging that is immediately compatible with downstream AI-based disease classification and can be readily deployed in conventional clinical ultrasound workflows-while maintaining 98% SSIM and 0.09% MSE in real time imaging frame rate, faster and more accurate than the previous. This focus on synthesizing Nakagami images for breast cancer diagnosis, rather than merely translating appearance between modalities or simulating segmentation labels, makes NakaSynthNet uniquely able to leverage the vast archive of clinical B-mode scans to produce diagnostically meaningful backscatter statistics on-the-fly for downstream CNN-based classification.

Having demonstrated real-time Nakagami map synthesis from B-mode inputs with NakaSynthNet that focuses on quantitative features of ultrasound images, SegmentNet leverages the B-mode images to accurately delineate tumor boundaries to maintain the qualitative features with the Nakagami quantitative features equally. Unlike early rule-based and level-set methods such as edge-based level set (EBLS) and level set-based active contour (LSAC)— which segment breast ultrasound lesions by thresholding and local gray-level statistics but suffer from high false-positive rates (SI = 70.93% for EBLS, SI = 80.33% for LSAC) and sensitivity to speckle noise— SegmentNet uses a lightweight Attention U-Net–derived encoder–decoder with skip connections and dropout to learn robust features directly from B-mode images [17]. This data-driven approach yields a Dice coefficient of 94% (compared to quasi-3D U-Net’s median Dice 78% on volumetric DCE-MRI slices) without requiring 3D inputs or contrast agents [19]. Moreover, whereas advanced attention-based CNNs incorporating CBAM or Non-Local Attention in recent ultrasound studies achieved Dice scores in the 60%–70% range with millions of parameters [22], SegmentNet attains 98.4% accuracy and only 0.4% BCE loss at 20 fps with a far more compact architecture. By focusing on direct 2D lesion delineation in routinely saved B-mode frames, SegmentNet is both substantially more accurate and faster than all models in these uploaded studies.

Muhtadi et al. channel-fused EfficientNetV2B0 requires precomputed Nakagami m and Ω maps—an extra manual step that increases computational overhead—and yields AUC = 87% by stacking B-mode, m, and Ω-maps as three channels [13], [30]. Their VGG-16 fusion model similarly relies on manually calculated Nakagami maps weighted against B-mode to achieve AUC = 87.1%, but lacks any explicit segmentation or qualitative features. In contrast, FeatureNet takes a single overlay image combining the NakaSynthNet-predicted Nakagami map and the SegmentNet-predicted lesion mask—both generated automatically from B-mode—thus obviating manual Nakagami computation and annotation. This dual quantitative (Nakagami) and qualitative (lesion mask) input allows FeatureNet to run in real time with minimal resources while achieving higher AUCs (98%) on benchmark datasets.

The simulated dataset was created to address the common problem of limited annotated clinical ultrasound images by synthesizing a diverse array of B-mode and Nakagami image pairs in silico. Rather than relying solely on scarce in vivo scans, the pipeline generates both natural-image–based and digital lesion phantom simulations under varying contrast, noise, and compression settings. This unique approach produces 110,247 of labeled B-mode/Nakagami pairs—each with known ground-truth parametric maps and lesion masks—without manual annotation. By exposing NakaSynthNet and SegmentNet to a wide spectrum of tissue appearances and acquisition artifacts, the simulated data substantially mitigates overfitting and enhances generalization, enabling robust training even when real clinical examples are few.

While our proposed framework achieved a sensitivity of 93% and specificity of 98%, comparable or better than existing performance, a small dataset is the main limitation of the study. However, this study lays the groundwork for synthesizing quantitative ultrasound parameters from B-mode ultrasound images and combining multimodal imaging with lesion masks for breast cancer. Future studies should investigate the synthesis of additional QUS parameters from B-mode images and integrate these parameters with lesion masks and explainable AI methodologies—while also expanding clinical datasets—to further enhance diagnostic precision and strengthen the clinical utility of this innovative approach.

## V. CONCLUSION

This study demonstrated an integrated deep learning framework combining real-time Nakagami parametric imaging, automated lesion segmentation, and robust classification using transfer learning, effectively enhancing the diagnostic accuracy of breast cancer diagnosis via ultrasound imaging. Besides proposing NakaSynth to generate Nakagami from B-mode and SegmentNet for automatic lesion segmentation, the proposed FeatureNet achieved an accuracy of 96.7%, a sensitivity of 93%, a specificity of 98%, and an AUC of 98% using a lesion mask overlaid on Nakagami parameters as input, compared to other input combinations. Real-time synthesis of Nakagami from B-mode makes it easily implementable in different ultrasound systems and applicable to various organs. The method to simulate B-mode images from natural and mammography images to augment scarce clinical ultrasound imaging datasets is helpful for other researchers developing AI-based methods. In conclusion, this integrated framework not only delivers substantial improvements in breast cancer detection accuracy but also establishes a scalable foundation for broad clinical adoption and cross-organ applications. By enabling efficient data augmentation and facilitating seamless integration with existing ultrasound systems, this approach paves the way for future advancements in AI-driven medical imaging and holds significant promise for improving patient outcomes across diverse settings.

## Data Availability

All human data used in this study are publicly available online, with details provided in the manuscript. The raw image data used to generate the synthetic datasets are also publicly available online. The methods for generating the synthetic data are fully described in the manuscript, and the resulting synthetic datasets are available from the corresponding author upon reasonable request.

https://scholar.cu.edu.eg/?q=afahmy/pages/dataset

https://bluebox.ippt.gov.pl/~hpiotrzk

https://data.mendeley.com/datasets/vkht8pfsp3/3

https://www.nature.com/articles/sdata2017177

## APPENDIX

Fig. 8 shows B-mode and Nakagami representation of natural images with scatterer intensities of –20 dB, –10 dB, +10 dB, and +20 dB at SNR levels of 5 dB, 15 dB, 25 dB, and 35 dB. These panels illustrate how variations in scatterer contrast and noise influence both the quantitative Nakagami estimates (color-coded) and the visual appearance of B-mode data under −50 dB and −80 dB log compression.

**Fig. 8.**
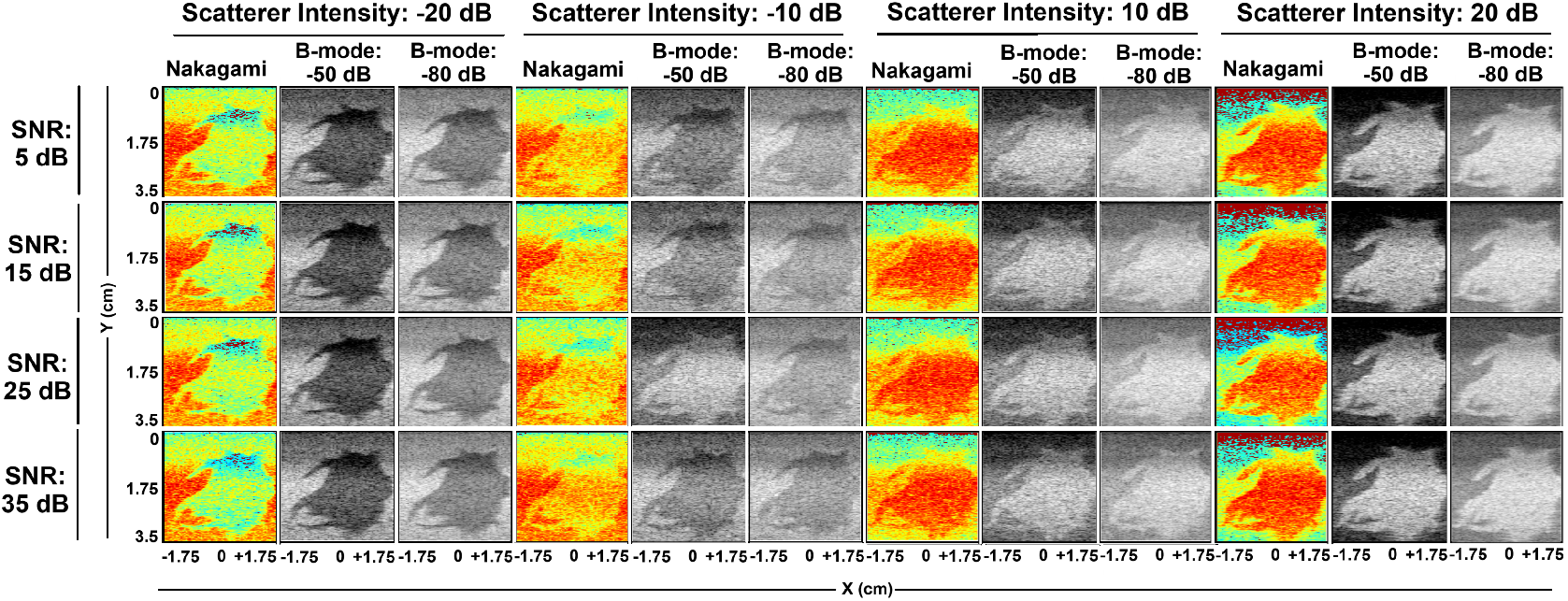
Representative calculated Nakagami and log-compressed B-mode images at varying scatterer intensities (–20 dB, –10 dB, +10 dB, +20 dB) and SNR levels (5 dB, 15 dB, 25 dB, 35 dB). Each column shows (left) the colorized Nakagami map, (center) the B-mode image with –50 dB log compression, and (right) the B-mode image with –80 dB log compression.

## Notes

### Competing Interest Statement

The authors have declared no competing interest.

### Funding Statement

This study was funded by Dr Hossain's startup award

### Author Declarations

W. Al Dhabyani, M. Gomaa, H. Khaled, and A. Fahmy, Dataset of breast ultrasound images, Data in Brief, vol. 28, 2 2020.H. Piotrzkowska Wroblewska, K. Dobruch Sobczak, M. Byra, and A. Nowicki, Open access database of raw ultrasonic signals acquired from malignant and benign breast lesions, Medical Physics, vol. 44, no. 11, 11 2017. R. S. Lee, F. Gimenez, A. Hoogi, K. K. Miyake, M. Gorovoy, and D. L. Rubin, A curated mammography data set for use in computer aided detection and diagnosis research, Scientific Data, vol. 4, no. 1, p. 170177, 12 2017.

